# Declined antibody responses to COVID-19 mRNA vaccine within first three months

**DOI:** 10.1101/2021.04.19.21255714

**Authors:** Paul Naaber, Liina Tserel, Kadri Kangro, Epp Sepp, Virge Jürjenson, Ainika Adamson, Liis Haljasmägi, Pauliina Rumm, Regina Maruste, Jaanika Kärner, Joachim M. Gerhold, Anu Planken, Mart Ustav, Kai Kisand, Pärt Peterson

**Author notes:** **Correspondence**: Paul Naaber, Veerenni 53a, 10138 Tallinn Estonia.

## Abstract

**Background:** Although the SARS-CoV-2 mRNA vaccines have proven high efficacy, limited data exists on the duration of immune responses and their relation to age and side effects.

**Methods:** We studied the antibody and memory T cell responses to Spike protein after the two-dose Comirnaty mRNA vaccine in 122 volunteers up to 3 months and correlated the findings with age and side effects.

**Findings:** We found a robust antibody response after the second vaccination dose. However, the antibody levels declined at 6 and 12 weeks postvaccination, indicating a waning of the immune response. Regardless, the average levels remained higher compared to pre-vaccination or in COVID-19 convalescent individuals. The antibodies efficiently blocked ACE2 receptor binding to Spike protein of four variants of concern at one week but this was decreased at three months, in particular with B.1.351 and P1 isolates. 87% of individuals developed Spike-specific memory T cell responses, which were lower in individuals with increased proportions of immunosenescent CD8+ TEMRA cells. We found a decreased vaccination efficacy but fewer adverse events in older individuals, suggesting a detrimental impact of age on outcome.

**Interpretation:** The mRNA vaccine induces a strong antibody response to four variants at 1 week postvaccination but decreases thereafter, in particular among older individuals. T cell responses, although detectable in the majority, were lower in individuals with immunosenescence. The deterioration of vaccine response needs to be monitored to define the optimal time for the revaccination.

**Funding:** The Estonian Research Council, Icosagen Cell Factory, and SYNLAB Estonia.

**Research in context:** *Evidence before this study:* The first studies addressing the immune responses in older individuals after the administration of SARS-CoV-2 mRNA vaccines have been published. We searched PubMed and medRxiv for publications on the immune response of SARS-CoV-2-mRNA vaccines, published in English, using the search terms “SARS-CoV-2”, “COVID-19”, “vaccine response”, “mRNA vaccine”, up to May 20th, 2021. To date, most mRNA vaccine response studies have not been peer-reviewed, and data on the dynamics of antibody response, role of age and side effects on SARS-CoV-2-mRNA vaccines in real vaccination situations is limited. Some studies have found a weaker immune response in older individuals after the first dose and these have been measured at a relatively short period (within one to two weeks) after the first dose but little longer-term evidence exists on the postvaccination antibody persistence.

*Added value of this study:* In this study, we assessed the antibody response up to three months after the full vaccination with Pfizer-BioNTech Comirnaty mRNA vaccine in 122 individuals. Our findings show strong Spike RBD antibody responses one week after the second dose with the capacity to block ACE2-Spike protein interaction, however, the antibodies declined significantly at three months after the second dose. The inhibition of ACE2-Spike interaction was weaker with South African (B.1.351) and Brazilian (P.1) than with Wuhan and UK (B.1.1.7) SARS-CoV-2 isolates. At three months 87% of vaccinated individuals developed either CD4+ or CD8+ T cell responses. Those negative for Spike-specific T cell response also tended to have lower Spike-specific antibody levels. In addition, CD4+ T cell response was decreased among vaccinated individuals with elevated levels of senescent CD8+ TEMRA cells. We found a weaker antibody response and faster waning of antibodies in older vaccinated individuals, which correlated with fewer side effects at the time of vaccinations.

*Implications of all the available evidence:* Our results show that two doses of Pfizer-BioNTech Comirnaty mRNA vaccine induce a strong antibody and T cell responses to Spike RBD region but the antibody levels are declined at three months after the second dose. Nevertheless, even at three months, the anti-Spike RBD antibody levels stay significantly higher than at prevaccination, after the first dose of vaccine, or in Covid-19 postinfection. Our findings implicate older individuals to have fewer vaccination adverse effects and weaker immune response after the vaccination and point to the need for more individualized vaccination protocols, in particular among older people.

## Introduction

New mRNA vaccines have shown high efficacy in clinical trials and are applied worldwide to millions of people. The first two-dose COVID-19 mRNA vaccine, Pfizer-BioNTech Comirnaty (BNT162b2), accepted for emergency use, was found safe and demonstrated 95% efficacy in phase 3 trials. However, so far exists little data about the extent and duration of the antibody and T cell responses after the mRNA vaccination, as well as about the factors influencing the efficacy and side effects in real vaccination situations.

Although highly efficient in triggering immune responses in clinical trials and initial real situation vaccinations, the first published short-term studies with Pfizer-BioNTech mRNA vaccines have reported weaker immune responses and a higher number of non-responders among older people after the two-dose vaccination with BNT162b2 vaccine^1-3^. Nevertheless, one study failed to show a significant correlation between age and antibody response after the second vaccination but found a lower magnitude of memory B cell responses with increased age ^4^ highlighting a need for further studies to understand the age-related responses to mRNA vaccination and to monitor for longer periods than less than one month. Also, limited information is available about the side effects and their correlation with vaccination outcomes. For example, one study found no significant association between the antibody levels and severity of adverse events among vaccinees^4^. Furthermore, few preprint studies have reported sex differences in response to COVID-19 vaccination^1,5^, although widely described with several other vaccines^6^. Here we addressed the dynamics of anti-S-RBD IgG and CD4+ and CD8+ T cell responses at three months after two doses of the Pfizer-BioNTech Comirnaty mRNA vaccine in healthy volunteers and assessed its correlation with the age and severity of side effects.

## Material and Methods

### Recruitment, sample, and data collection

SYNLAB Estonia employees volunteering to be vaccinated with COVID-19 mRNA Comirnaty (Pfizer-BioNTech) vaccine were invited to participate in the study. Participants signed an informed consent form agreeing with sampling and usage of their clinical data. The blood samplings were performed by trained medical personnel at SYNLAB Estonia. Two doses were given three weeks apart, and the samples were taken before the first dose of vaccine (B1D), before the second dose (B2D), one week after the second dose (1wA2D), six weeks after the second dose (6wA2D), and 12 weeks after the second dose (12wA2D). The study participants filled in a questionnaire about the presence of side-effects after the second dose and rated their side-effect severity with scoring from zero to three (**Supplementary Table 1**). All samples and volunteers’ data (age, sex, side effects) were stored in a pseudonymized manner. As controls, we used samples from uninfected and non-vaccinated negative controls (n=50) and PCR-positive mild COVID-19 (n=97) patients collected and described previously^7^.

The study has been approved by the Research Ethics Committee of the University of Tartu on February 15, 2021 (No 335/T-21). Patients signed informed consent before recruitment into the study.

### Antibody testing

Serum samples were analysed for the IgG antibodies to SARS-CoV-2 Spike protein receptor-binding domain (S-RBD) IgG using quantitative Abbott SARS-CoV-2 IgG QN (all time points) and anti-Spike IgM using Abbott SARS-CoV-2 IgM (B1D, B2D, 1wA2D). In both analyses, chemiluminescent micro-particle immunoassay (CLIA) on ARCHITECT i2000SR analyser (Abbott Laboratories) was applied. The cut-off and the upper detection limit of the IgG test were 50 and 80,000 AU/mL, respectively. The results of S-RBD IgM were interpreted as positive or negative by the analyser.

### ACE2-Spike interaction blocking assay

The serum capacity to block the angiotensin-converting enzyme 2 (ACE2) receptor interaction with SARS-CoV-2 trimeric S protein receptor-binding domain (RBD) was tested using IVD-CE SARS-CoV-2 Neutralizing Antibody ELISA kit (Icosagen). In brief, the ELISA plates covered with SARS-CoV-2 trimeric S proteins of Wuhan, UK (B.1.1.7), South African (B.1.351), and Brazilian (P.1) isolates (Icosagen) were incubated with serum samples in a 1/100 dilution and probed with biotinylated ACE2-hFc protein (Icosagen). Streptavidin HRP was used for colorimetric detection and the light absorbance was measured at 450 nm as optical density (OD) values. The OD values of the measured samples were divided by the mean value of the three repeats of NC to obtain relative OD values. The samples with relative OD values of <0·75 were considered sufficient in blocking ACE2 binding.

### SARS-CoV-2 Spike-specific CD4+ and CD8+ memory T cell responses

For CD4+ and CD8+ T cell response analysis, freshly isolated PBMCs (2×10^6^ cells) were stimulated with overlapping SARS-CoV2 S peptide pool (1ug/ml, Miltenyi Biotec), and with anti-CD28 and anti-CD49d for 20 hours. CEFX peptides (JPT Peptides) were used as a positive control. After the stimulation T cells were stained for CD3 Brilliant Violet 650, CD4 Alexa Fluor 700, CD8 Brilliant Violet 605, CCR7 Alexa Fluor 488, CD45RA APC, CD69 Brilliant Violet 510, OX40 PE-Dazzle (all from Biolegend), and CD137 PE (from Miltenyi Biotech). 7AAD was used for the discrimination of dead cells. Flow cytometry was performed using LSRFortessa (BD Biosciences) and the results were analysed with FCS Express 7 (DeNovo Software). Antigen-specific cells were gated according to the upregulation of activation-induced markers (AIM) CD137 and CD69 in memory CD8+ T cells and CD137, OX40, and CD69 in memory CD4+ T cells. The percentage of AIM positive cells in the negative control sample (diluent with costimulatory antibodies) was subtracted from the value from the stimulated sample. The cut-off level of positivity was drawn to 0·02% according to the data from six unvaccinated individuals.

### Statistics

The t-test, paired t-test, and Pearson correlation calculations were used for data analysis using PAST 4.05 software. GraphPad version 9 was used to do statistical analyses of Kruskall-Wallis tests.

## Results

We studied 122 individuals (21 males and 101 females) who received their first and second COVID-19 Pfizer-BioNTech vaccine doses and gave corresponding pre- and post-vaccination blood samples. From these, 90 completed the questionnaire on post-vaccination side effects. The age of the vaccinated volunteers ranged from 21 to 69 years (median 37). The number of participants in each analysis is presented in **Table 1**.

**Table 1.**
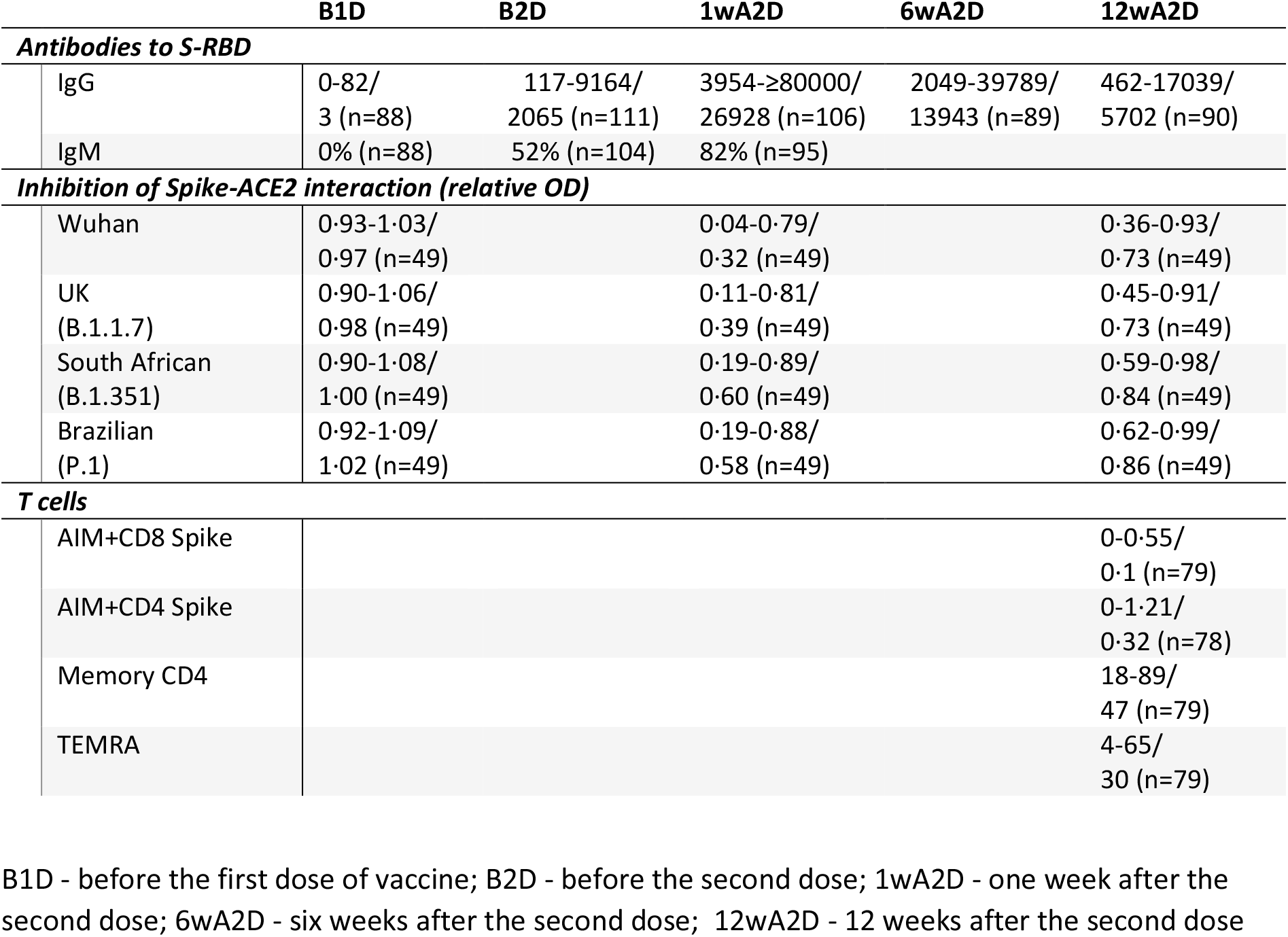
Summary statistics of each analysis, giving the range, mean, and number of studied individuals. For S-RDB IgM antibodies, the percentages of positive individuals are shown.

### Antibody dynamics

Three weeks after the first vaccine dose, we found elevated S-RBD IgG levels in vaccinated serum samples (**Figure 1**), measured by the Abbot Laboratories CLIA method, with mean IgG levels of 2079 AU/mL (**Table 1**). Importantly, these S-RBD IgG levels increased significantly after the second vaccination dose (at 1 and 6 weeks) as compared to the first dose (p<0·0001). One week after the second dose, the serum antibody levels were boosted in participants to a mean level of 26928 AU/mL. However, we found that the S-RBD IgG levels were significantly decreased to 13943 AU/mL (p<0·0001) at six weeks and to 5702 AU/mL at 12 weeks after the second dose as compared with previous time points (**Figure 1**). The dynamics of declining antibody levels between one and six weeks after the second dose was present in most of the vaccinees, and on average S-RBD IgG levels decreased 45% between these two time-points **(Supplementary Figure 1)**. In contrast, we found increased S-RBD IgG levels at six weeks after the second dose in only four % of individuals. The further decline of the S-RBD IgG levels at 12 weeks after the second dose was present in all participants, and at this time point, the S-RBD IgG levels were up to only 23% of their peak levels, detected at one week after the second dose.

**Figure 1.**
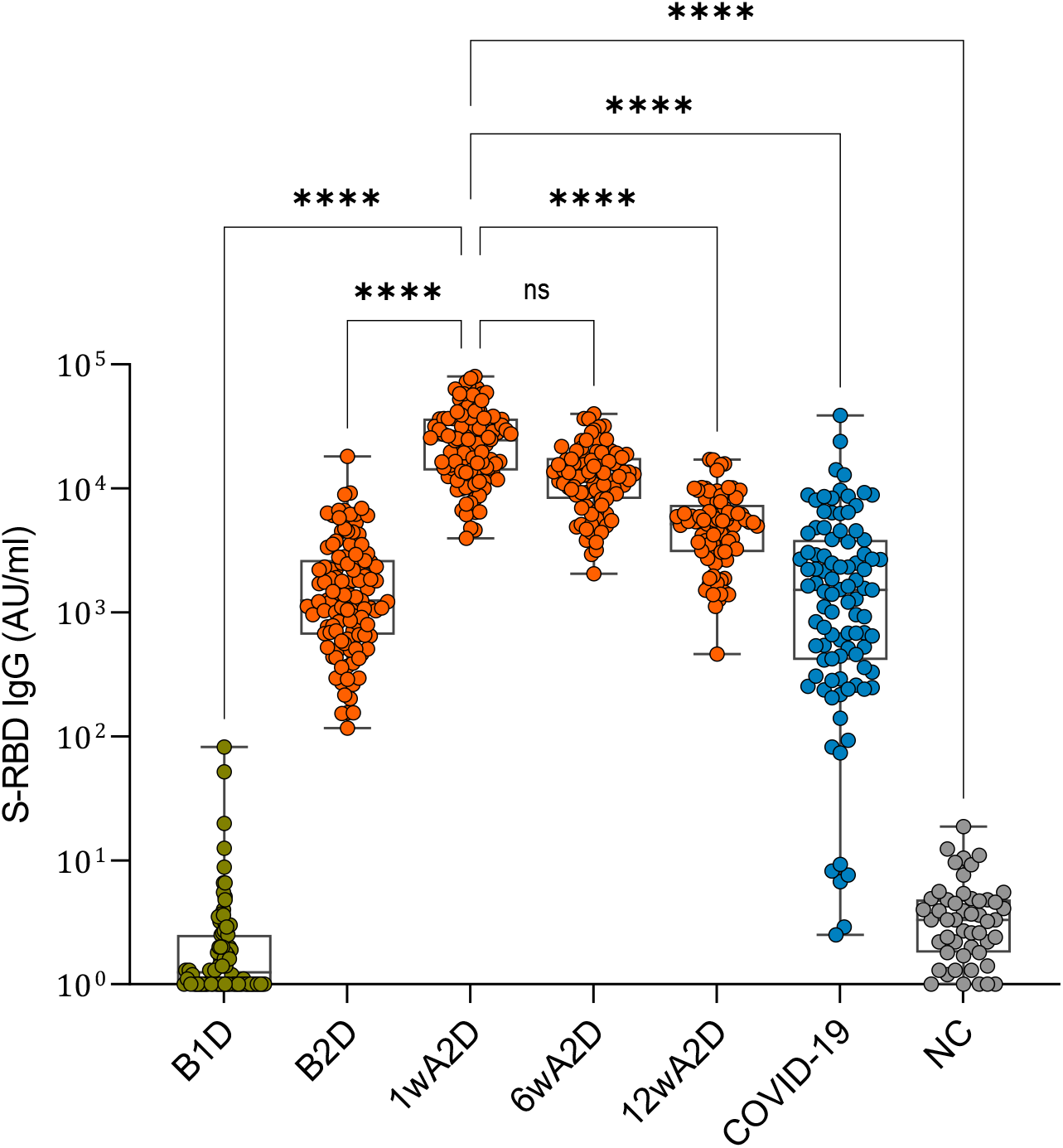
S-RBD IgG levels before vaccination, after the single and two-dose immunizations with Pfizer-BioNTech vaccine compared with pre-COVID-19 negative controls and post-infection levels in patients recovered from COVID-19. The box plot comparisons of the S-RBD IgG levels in vaccinated individuals after the first vaccine dose, one, six and 12 weeks after the second dose, and in COVID-19 convalescent and negative control sera (Kruskall-Wallis test **** p<0·0001).

One individual had slightly elevated S-RBD IgG before vaccination, but negative anti-Nucleocapsid IgG and anti-Spike IgM, also the post-vaccination S-RBD IgG was close to average. Although COVID-19 was never diagnosed in this person we could not exclude possible earlier exposure to SARS-CoV-2. We also saw a greater S-RBD IgG decrease in individuals with higher IgG values a week after the second dose (r= 0·27, p=0·02).

IgM was negative in all tested pre-vaccination samples, positive in 52% of B2D, and 82% of 1wA2D samples. In IgM-positive persons, the IgG levels were significantly higher as compared with IgM negative ones.

We also compared the post-vaccination results with S-RBD antibodies in COVID-19 recovered patients (**Figure 1**). The post-infection IgG levels (mean 3932) were somewhat higher than in vaccinated persons who received the first dose (p=0·01) but were significantly lower than in those who received two vaccine doses (p<0·0001).

### Inhibition of ACE2-trimeric Spike interaction by vaccine-induced antibodies

We next tested the inhibition of ACE2 interaction with trimeric S protein from SARS-CoV-2 Wuhan, UK, South African, and Brazilian isolates by the sera of vaccinated participants. The serum samples collected before the vaccination did not block ACE2 binding to trimeric S protein of any of the isolates analyzed (**Figure 2**). In contrast, all serum samples collected one week after the second dose (1wA2D) were able to block this interaction with average relative OD values at 0·6, albeit this was less pronounced with the trimeric S variants of UK (B.1.1.7), South African (B.1.351) and Brazilian (P.1) isolates as compared with Wuhan variant p<0·00001). However, 12 weeks after the administration of the second dose (12wA2D), we found diminished values of relative OD ranging around the set threshold of 0·75 with the Wuhan and UK (B.1.1.7) isolates, while the blocking capacity waned even more with South African (B.1.351; p<0·00001) and Brazilian (P.1; p<0·00001) isolates. Following S-RBD IgG increase after the vaccination, we found strong correlations between ACE2-trimeric S blocking capacity, with all isolates, and S-RBD IgG levels in 1wA2D and 12wA2D groups (p<0·00001; r= from −0·84 to −0·9; **Supplementary Figure 2 and 3**). The results show that post-vaccination, the blocking antibodies peak at one week after the second dose but then decline, as seen 12 weeks after the second dose. Our findings also indicate a strong correlation between S-RBD interacting and RBD inhibiting IgG levels.

**Figure 2.**
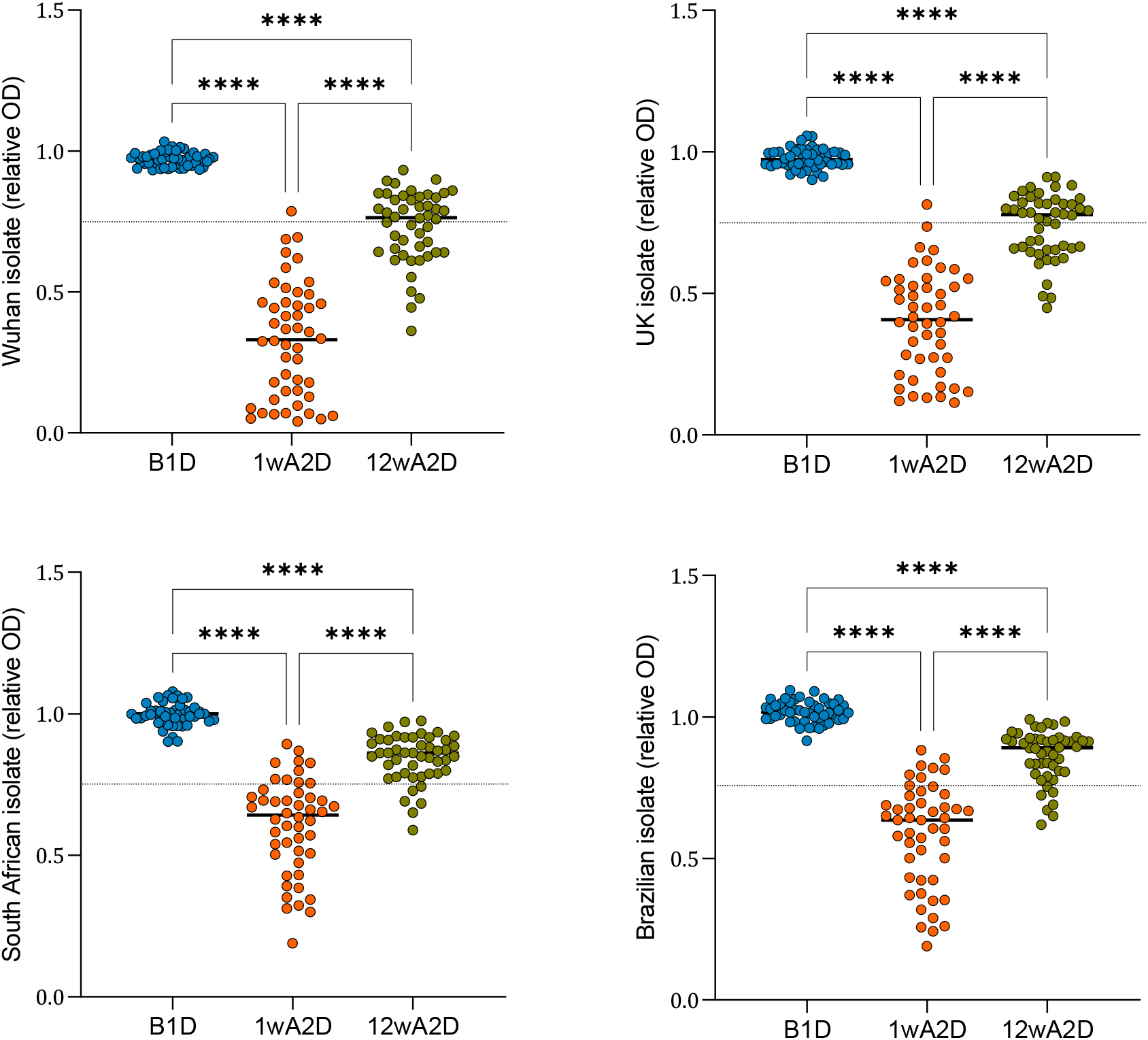
Inhibition of ACE2-trimeric Spike interaction by vaccine-induced antibodies. Serum antibody capacities to block the interaction of ACE2 receptor and Spike protein with the modifications of four different isolates, Wuhan, UK, South African, and Brazilian were analyzed before the vaccination, one and 12 weeks after the second dose (Kruskal-Wallis test **** p<0·0001). The dotted line indicates the relative OD value of 0·75, which is a threshold for sufficient blocking of ACE2 binding.

### T cell responses

We found 74% and 73% of vaccinated individuals to have CD4+ and CD8+ memory responses, respectively, to Spike peptide pools measured by upregulation of CD69, OX40, and CD137 activation markers, indicating that the majority of vaccinated individuals developed SARS-CoV-2 - specific memory T cell responses 12 weeks after the second dose. Collectively, 87% of vaccinated individuals developed either CD4+ or CD8+ T cell responses to the vaccine. The vaccinated individuals who did not develop T cell responses after the three months also had slightly lower S-RBD antibody responses (**Figure 3A**). We next analysed the proportions of T cell subsets, in particular immunosenescence-associated CD8+ TEMRA population, as measured by CCR7 and CD45RA markers in vaccinated individuals. Interestingly we found that individuals with the higher frequency of CD8+ TEMRA cells developed lower cell-mediated responses to immunized Spike protein (**Figure 3B**), suggesting that T cell-related immunosenescence has a detrimental effect on the development of cell-mediated protection to the SARS-CoV-2 virus.

**Figure 3.**
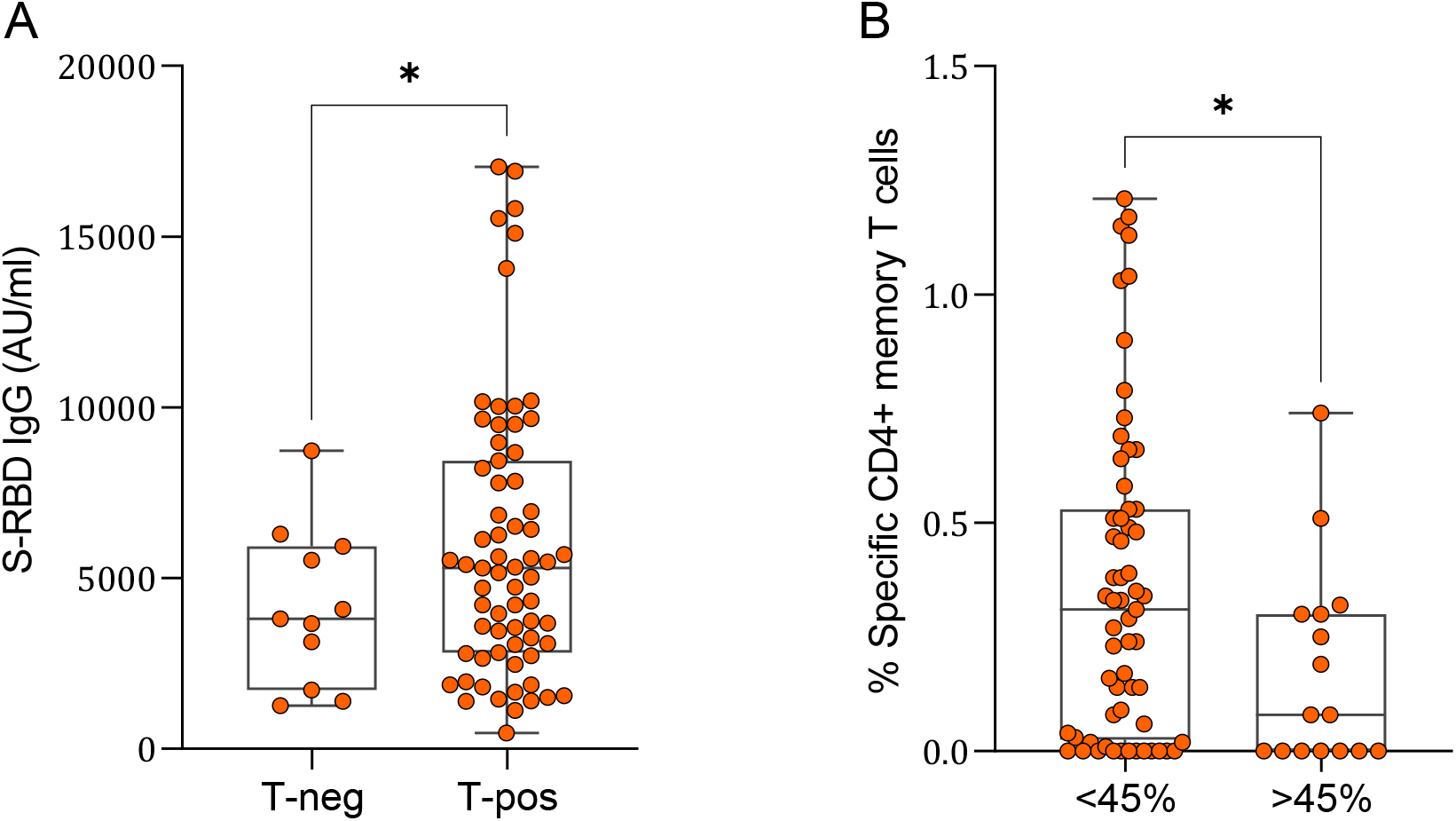
Spike-specific T cell responses in comparison with S-RBD antibodies and the proportions of CD8+ TEMRA cells. (A) S-RBD IgG values in vaccinated individuals with (T-pos) or without (T-neg) S-specific T cell responses. (B) Post-vaccination S-specific memory CD4+ T-cells in individuals with low (<45%) and high (>45%) percentage of CD8+ TEMRA cells (two-tailed t-test * p < 0·05).

### Factors influencing the vaccination response

The age of vaccinated individuals had a significant negative correlation with S-RBD IgG response in all measured time points. This was the strongest at B2D timepoint (r= −0·41, p<0·0001), but less significant, although still detectable, at 1wA2D (r= −0·34, p<0·001), 6wA2D (r= −0·25, p=0·02), and 12wA2D (r= −0·3, p=0·003); **Supplementary Figure 2**). We found a similar association while comparing the antibody levels in persons either younger or older than 40 years. The individuals less than 40 years had higher antibody levels after the first dose (means 2732 vs 1147 AU/mL; p=0·001), as well as 1 week (30375 vs 19934 AU/mL; p=0·003) and 12 weeks after the second dose (6386 vs 4520 AU/mL; p=0·022 **Supplementary Figure 4**).

In a comparison of sex differences, we found a stronger association between age and immune response among males, where the negative correlations between age and S-RBD antibodies were equally strong until six weeks after the second dose - r=-0·58 (B1D; p=0·01), r=-0·59 (1wA2D; p=0·01), and r=-0·58 (6wA2D; p<0·05). In females, the corresponding correlations were weaker and also declined in time r= −0·37 (p<0·001), r=-0·28 (p=0·01), and r=-0·20 (ns), suggesting that age together with sex-specific factors affect the vaccination outcomes of mRNA vaccines.

### Side effects of mRNA vaccination

Vaccination side-effects merit investigation as they are common reasons for vaccine hesitancy. Altogether 93% of participants reported some type of adverse effects. The most common side effects were reported pain or swelling (in 84%) at the injection site, fatigue (64%), malaise (50%), headache (42%), chills (41%), fever, and myalgia (both 34%). The majority of the side effects were present as mild to moderate. However, 20 (22%) persons reported one or several symptoms to significantly disturb daily life activities, and lasting for several days and/or causing absence from work. The total score of side effects (sum of all self-rated side effect scores per patient) ranged between zero and 27 (mean 7·76). The detailed data on individuals’ side effects are presented in **Supplementary Table 1**.

### Factors influencing the vaccine side effects

We found several side effects to positively correlate with the immune response to S-RBD. This was seen with the total score of adverse effects, which significantly associated with the S-RBD IgG levels at all time points i.e. B2D (r= 0·3, p<0·01), 1wA2D (r= 0·47, p<0·0001), 6wA2D (r= 0·46, p<0·0001), and 12wA2D (r= 0·41, p<0·001). An even stronger correlation was present with fever at all time-points: r= 0·36 (p<0·001), r= 0·47 (p<0·0001), r= 0·51 (p<0·0001), r= 0·49 (p<0·0001), and also other adverse symptoms such as headache, fatigue, malaise, chills, and nausea were positively correlated with the vaccine response (**Supplementary Figure 5**).

The age of vaccinated individuals negatively correlated with the total score of side effects (r= −0·35, p<0·001) as well as with several specific side effects (**Supplementary Figure 2**). Similar to the overall vaccination response, when comparing the two sexes separately, we again found males to have a stronger correlation of vaccine side effects with S-RBD antibody levels. The associations between the score of all side effects and IgG levels were strikingly higher in males than in females at one week after the second dose r= 0·87 (p<0·0005) vs r= 0·44 (p<0·0001) and at six weeks after the second dose r= 0·72 (p<0·01) vs r= 0·44 (p<0·001). Together the results indicate age- and sex-related variations in side effects to mRNA vaccination responses.

## Discussion

We report S-RBD IgG responses after COVID-19 mRNA vaccination showing a significant initial increase in antibody levels after the second dose. However, at three months post-vaccination these levels were decreased to 23% on average of their peak level but they stayed significantly higher compared to patients recovered from COVID-19.

The vaccinated sera robustly inhibited the ACE2-Spike protein-protein interaction suggesting efficient induction of neutralizing antibodies by mRNA vaccination. The new SARS-CoV-2 variants with mutated structural properties of the spike glycoprotein have posed concern about their transmissibility, virulence, and neutralizing antibody escape. We here show that Comirnaty mRNA vaccine can generate neutralizing antibodies to all four isolates with slightly higher efficiency to Wuhan and UK (B.1.1.7) than to South African (B.1.351) and Brazilian (P.1) variants, confirming a recent report on BNT162b2-elicited neutralization against SARS-CoV-2 Spike of different variants^8^. At three months post-vaccination the neutralization capacity is significantly decreased, in agreement with lower S-RBD antibody levels. Previous studies have shown a decrease of neutralizing SARS-CoV-2 antibody levels to nucleocapsid within the first months after infection, especially in patients with mild COVID-19 (reviewed in ^9^). Currently, the long-lasting effect of mRNA vaccines to protect against reinfections or severe COVID-19 disease remains unclear, and might not only depend on antibody responses but also T cell immunity that we studied here. Nevertheless, our results highlight the importance of monitoring the antibody responses in vaccinated individuals to identify the need and time for potential revaccination.

The majority of the vaccinated individuals developed T cell responses with similar prevalence in both T cell subsets (74% for CD4+ and 73% for CD8+), in agreement with phase I/II clinical trials with mRNA vaccines, which have demonstrated activation of CD4+ and CD8+ T cells ^10,11^. Induction of functional T cells occurs after the BNT162b2 vaccination but with variable results in aged individuals, for example, Spike-specific IFNγ T cell responses to vaccines were impaired in the over 80 age group ^12^. Our findings showed that individuals with the increased number of immunosenescent CD8+ TEMRA cells had a lower number of Spike-specific T cell responses. This suggests immunosenescence, impairment of immune response to pathogens and vaccines, may affect vaccine response to SARS-CoV-2 and pose significant health challenges in vaccination of older individuals or those with comorbidities.

We found a negative correlation between antibody responses and the age of vaccinated individuals. Age is an important factor that influences vaccine responses, and elderly people have been reported to be poor responders to influenza, hepatitis A and B, and pneumococcal vaccines by developing lower antibody levels and weaker cell-mediated responses^6^. In addition to diminished post-vaccine responses, older individuals had a more rapid waning of antibodies after the vaccinations. The adverse effect of age on COVID-19 mRNA vaccination has been reported by other studies^1-3^. We here show weaker mRNA vaccine response after the first dose and also one, six, and 12 weeks after the second dose, confirming the previous results but also show that this difference equalizes and is less significant at six and 12 weeks after the second dose. Thus, our results indicate the benefit of the second dose or extended interval between the two doses^13^ for older individuals and its effect to level up the short-term vaccination response in younger and older persons, although the long-term persistence of post-vaccination antibody levels in older populations remains to be studied.

Interestingly, although we could not see direct sex differences in the vaccine response, we found a stronger negative association of age and S-RBD antibodies among males, indicating sex differences in mRNA vaccination, which has not been reported by other COVID-19 mRNA vaccination studies. The findings across the vaccination studies investigating the effect of sex on immune responses are largely consistent but not uniform with females having higher antibody responses to dengue, hepatitis A and B, inactivated polio, rabies, smallpox, and trivalent inactivated influenza vaccination, with males having higher antibody responses to diphtheria, meningococcal, pneumococcal, and tetanus vaccination^6^. Our findings show that age remains a negative factor for mRNA vaccination in males whereas in females the second dose, by and large, abolishes the detrimental impact of age on mRNA vaccination outcome. Despite the smaller representation of male participants in our study, this suggests less efficient mRNA vaccination response in older males and highlights the need to stratify vaccine responses among the elderly.

Common systemic side effects reported for COVID-19 mRNA vaccines are fatigue, headache, muscle pain, chills, and fever. In our study, 93% of vaccinated individuals reported some type of side-effects, which is higher than previously reported 66% of vaccinated seronegative persons^14^. In agreement with our results, the side effects among some groups were seen in 100% of participants of the mRNA vaccine phase 1/2 study^10^. An expected, older participants reported fewer or even no side effects as also seen earlier ^4^, and the presence and score of side effects correlated with S-RBD IgG responses.

Taken together we report a robust vaccine response after two doses of Pfizer-BioNTech Comirnaty (BNT162b2) vaccine with lower responses and fewer side effects in older individuals.

## Data Availability

Data is available on request

## Acknowledgments

We thank David James (SYNLAB UK) for language corrections. The study was supported by the Centre of Excellence in Translational Genomics (EXCEGEN), and the Estonian Research Council grant PRG377, PRG1117, Icosagen Cell Factory, and SYNLAB Estonia.

