## Supplementary for "Declined antibody responses to COVID-19 mRNA vaccine within first three months"

### Supplementary Material

**Supplementary Table 1. Prevalence of side effects in participants (n=90) after the second dose of Comirnaty vaccine and severity score**

| Side effects | % of participants | | | |
|---|---|---|---|---|
| | **Any score** | **Score 1*** | **Score 2** | **Score 3** |
| Injection site pain or swelling | 84 | 56 | 26 | 3 |
| Injection site redness | 20 | 18 | 2 | 0 |
| Injection site pruritus | 18 | 14 | 3 | 0 |
| Headache | 42 | 12 | 22 | 8 |
| Fatigue | 64 | 20 | 31 | 13 |
| Malaise | 50 | 13 | 24 | 12 |
| Chills | 41 | 14 | 22 | 4 |
| Fever | 34 | 12 | 17 | 6 |
| Myalgia | 34 | 10 | 20 | 4 |
| Arthralgia | 21 | 7 | 12 | 2 |
| Pain in extremity | 13 | 3 | 8 | 2 |
| Insomnia | 12 | 6 | 7 | 0 |
| Nausea | 11 | 6 | 4 | 1 |
| Diarrhea | 3 | 3 | 0 | 0 |
| Lymphadenopathy | 8 | 4 | 3 | 0 |
| Facial sensitivity disorder | 2 | 2 | 0 | 0 |
| Hypersensitivity/allergic reaction | 0 | 0 | 0 | 0 |

*Score: 1- mild symptoms that did not disturb daily life; 2- moderate symptoms that somewhat disturbed the activities of daily living; 3- symptoms that last days and/or caused absence from work

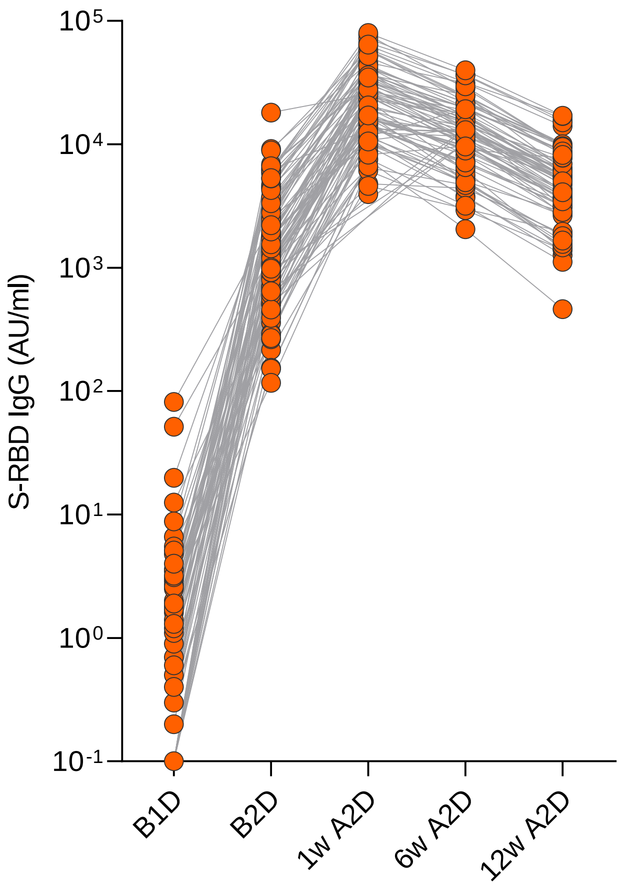


**Supplemental Figure 1. Line chart to show the results of S-RBD IgG antibody levels and dynamics between the sample values before the vaccination (B1D), before the second dose (B2D), and 1 (1wA2D), 6 (6wA2D) and 12 (12wA2D) weeks after the second dose.**

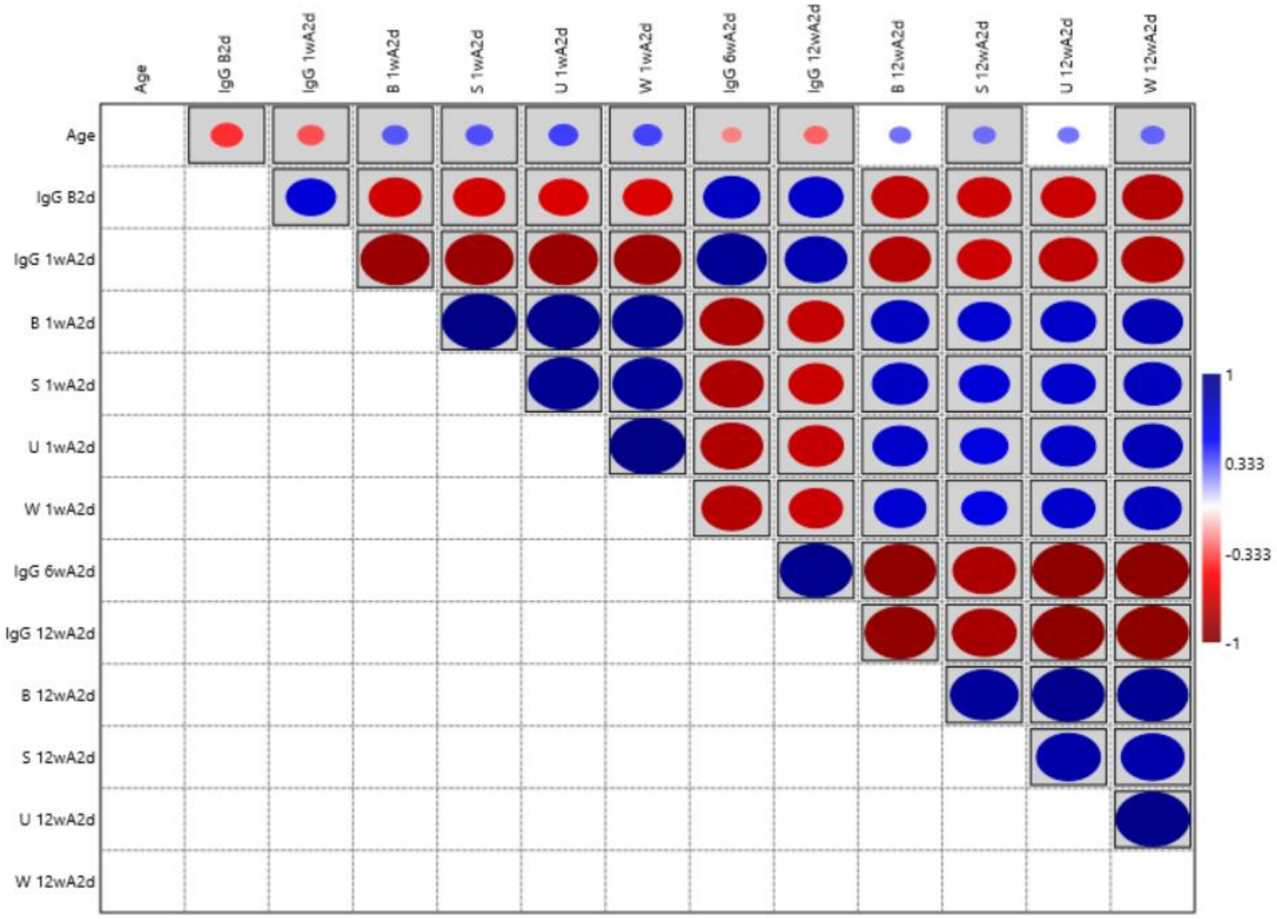


**Supplementary Figure 2. Correlations between age, S-RBD IgG levels and ACE2-trimeric S blocking capacity.** Higher size and intensity of dots indicates stronger correlation; Blue dots – positive correlation; Red dots – negative correlation; Boxed dots – statistically significant correlation ($p<0{\cdot}05$). W - Wuhan SARS-CoV-2 variant; U – UK variant (B.1.1.7); S – South African variant (B.1.351); B – Brazilian variant (P.1)

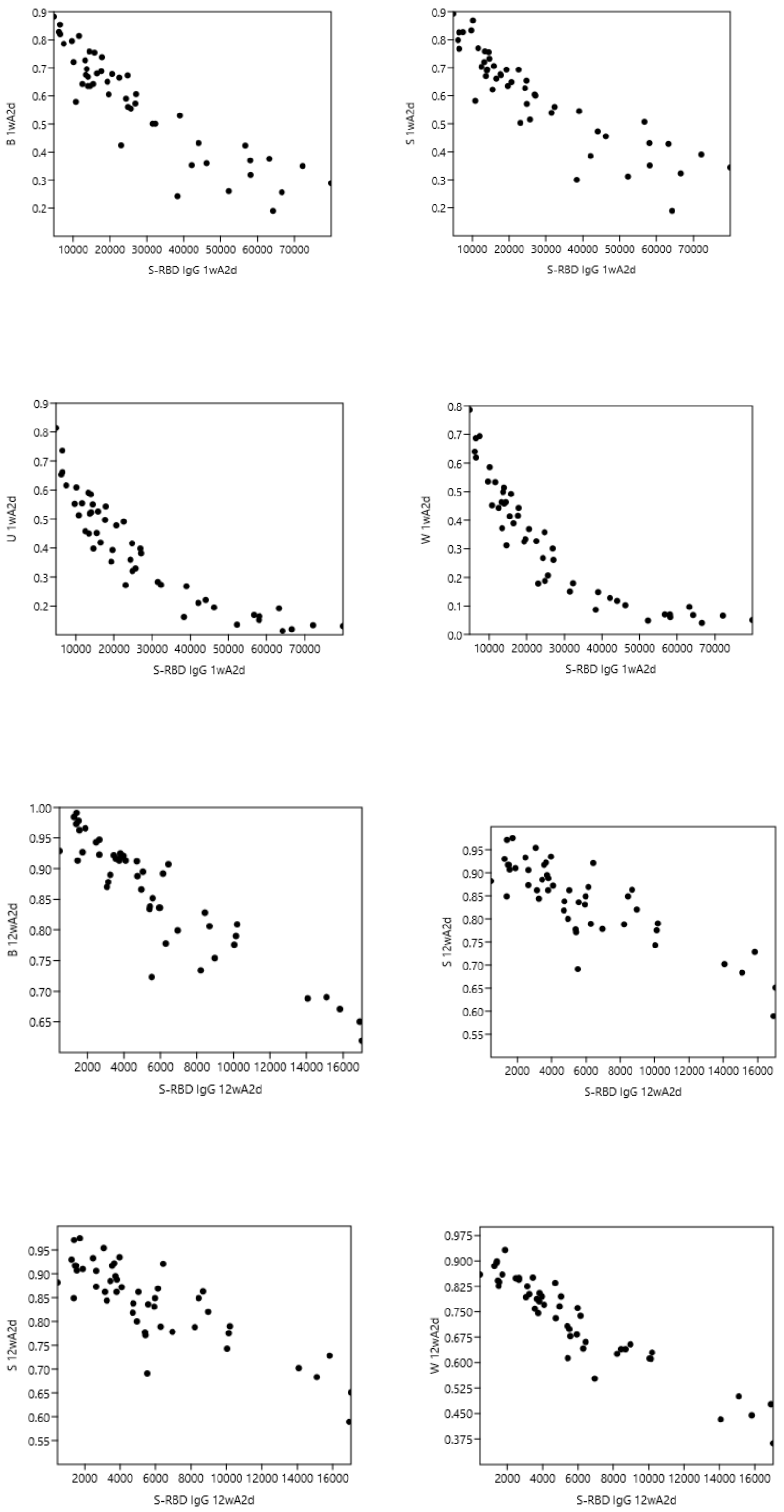


**Supplementary Figure 3. S-RBD IgG (AU/ml) and ACE2-trimeric S blocking capacity (relative OD) correlation blots at timepoints 1wA2A and 12wA2D.** W - Wuhan SARS-CoV-2 variant; U – UK variant (B.1.1.7); S – South African variant (B.1.351); B – Brazilian variant (P.1)

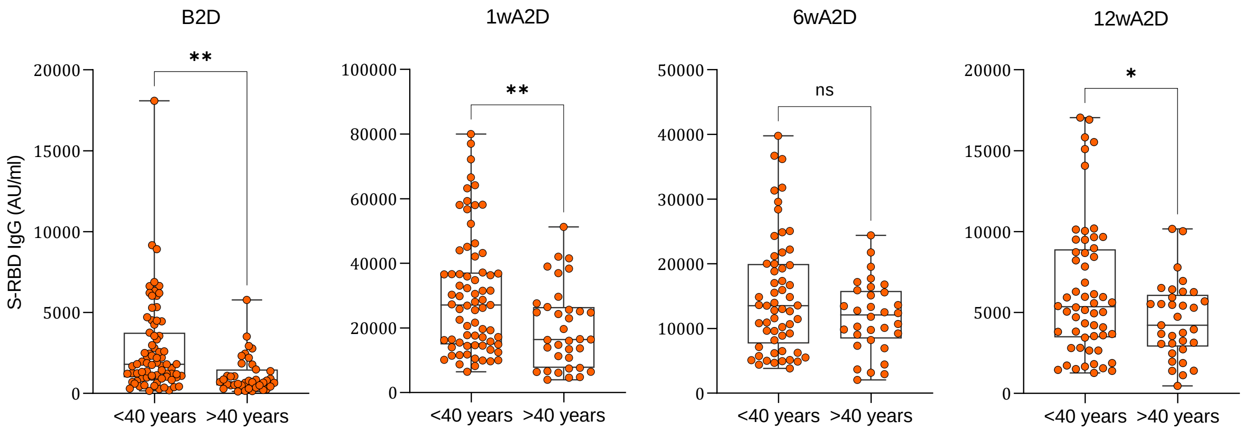


**Supplementary Figure 4. S-RBD IgG levels among persons >40 and ≥40 years old.** The group-wise box-plot comparisons of the S-RBD IgG levels in <40 and ≥40 years old vaccinated individuals after the first vaccine dose, and 1, 6, 12 weeks after the second vaccine dose (two-tailed t-test, ** p<0·001, * p<0·05).

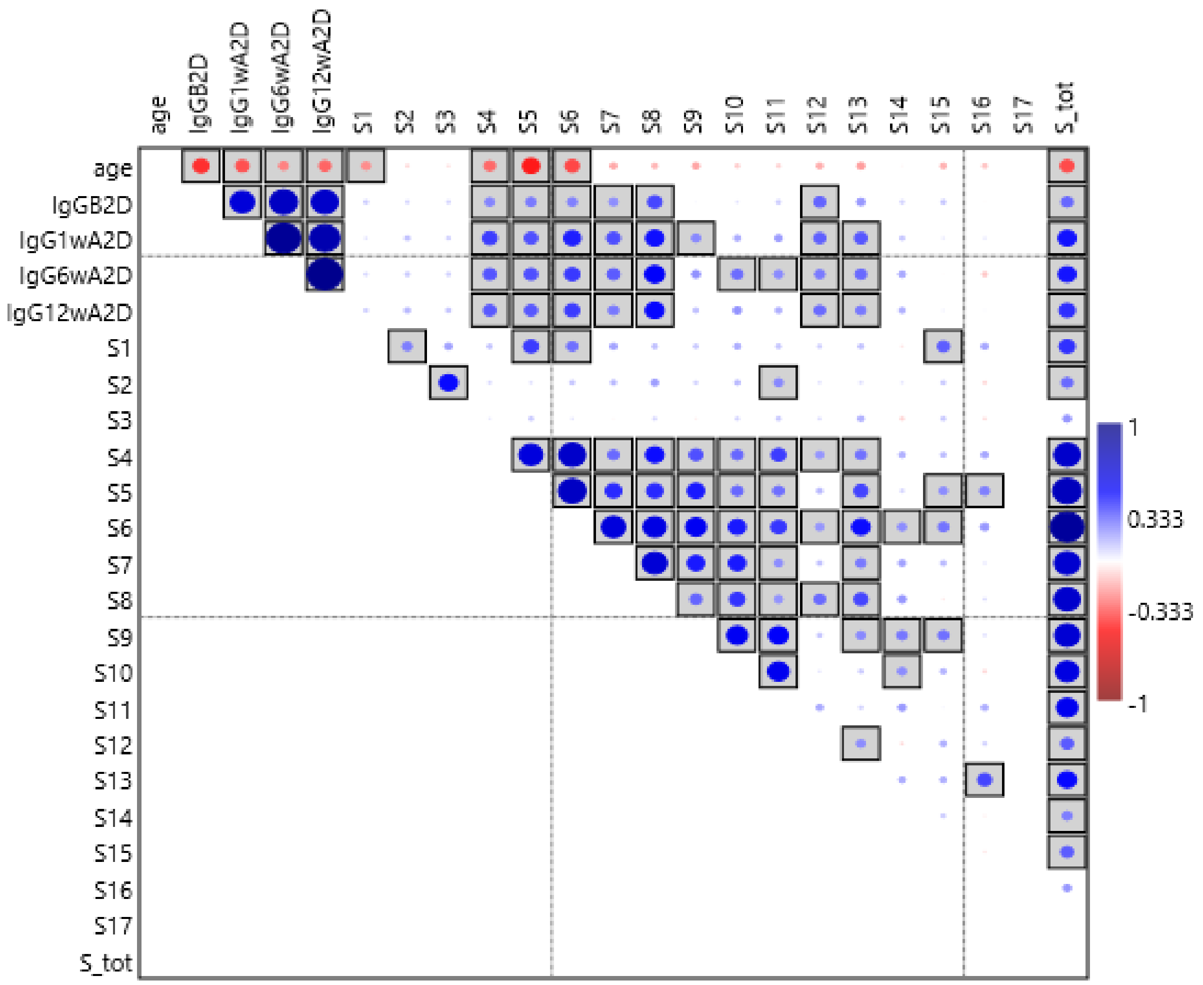


**Supplementary Figure 5. Correlations between age, S-RBD IgG levels and vaccine side-effects.** Higher size and intensity of dots indicates stronger correlation; Blue dots – positive correlation; Red dots – negative correlation; Boxed dots – statistically significant correlation ($p<0·05$)**.** Side effects: S1 - Injection site pain or swelling; S2 - Injection site redness; S3 - Injection site pruritus; S4 – Headache; S5 – Fatigue; S6 – Malaise; S7 – Chills; S8 – Fever; S9 – Myalgia; S10 – Arthralgia; S11 - Pain in extremity; S12 – Insomnia; S13 – Nausea; S14 – Diarrhea; S15 – Lymphadenopathy; S16 - Facial sensitivity disorder; S17 - Hypersensitivity/allergic reaction; S_tot – Total side effects score
